# Normative modeling of thalamic nuclear volumes

**DOI:** 10.1101/2024.03.06.24303871

**Authors:** Taylor Young, Vinod Jangid Kumar, Manojkumar Saranathan

## Abstract

Thalamic nuclei have been implicated in neurodegenerative and neuropsychiatric disorders. Normative models for thalamic nuclear volumes have not been proposed thus far. The aim of this work was to establish normative models of thalamic nuclear volumes and subsequently investigate changes in thalamic nuclei in cognitive and psychiatric disorders. Volumes of the bilateral thalami and 12 nuclear regions were generated from T1 MRI data using a novel segmentation method (HIPS-THOMAS) in healthy control subjects (n=2374) and non-control subjects (n=695) with early and late mild cognitive impairment (EMCI, LMCI), Alzheimer’s disease (AD), Early psychosis and Schizophrenia, Bipolar disorder, and Attention deficit hyperactivity disorder. Three different normative modelling methods were evaluated while controlling for sex, intracranial volume, and site. Z-scores and extreme z-score deviations were calculated and compared across phenotypes. GAMLSS models performed the best. Statistically significant shifts in z-score distributions consistent with atrophy were observed for most phenotypes. Shifts of progressively increasing magnitude were observed bilaterally from EMCI to AD with larger shifts in the left thalamic regions. Heterogeneous shifts were observed in psychiatric diagnoses with a predilection for the right thalamic regions. Here we present the first normative models of thalamic nuclear volumes and highlight their utility in evaluating heterogenous disorders such as Schizophrenia.

## Introduction

The thalamus is a bilateral subcortical structure with widespread connectivity to the cortex and subcortical structures including basal ganglia, brainstem, and cerebellum (Biesbroek et al. 2023). It serves as a relay center for primary sensory input to the cortex, facilitates movement and motor functional processing, and participates in higher cortical functions such as arousal, executive function, learning, memory, emotion, motivation, language, and multisensory integration (Schmahmann 2003). The thalamus and, specifically, its component nuclei have been implicated in several neurological, neuropsychiatric, and neurodegenerative conditions including essential tremor, multiple sclerosis, epilepsy, obsessive-compulsive disorder, schizophrenia, chronic pain syndrome, Alzheimer’s disease, and frontotemporal dementia (Neudorfer et al. 2024; Fujimori and Nakashima 2024; Burdette, Patra, and Johnson 2024; Yang et al. 2024; Alemán-Gómez et al. 2023; Wang et al. 2023; Forno et al. 2023; McKenna et al. 2023; Bernstein et al. 2021; Low et al. 2019). Until recently, the thalamus has been considered as a “whole” in most neuroimaging studies. This is primarily due to the lack of readily available tools for accurate segmentation and volumetry of the component thalamic nuclei from routine structural MRI. Specialized methods using advanced diffusion MRI and resting state functional MRI have been proposed to segment thalamic nuclei but are limited by poor spatial resolution, distortion, and more importantly, divergence from histological parcellation (Mastropasqua et al. 2015).

Very few studies have documented volumetric changes in thalamic nuclei with aging. One study (Hughes et al. 2012) characterized changes in thalamic shape with aging in 86 healthy volunteers. Two recent studies looked at thalamic nuclear volume changes as a function of age in healthy subjects using a relatively small number of subjects (198 and 237 respectively) from one or two sites (Choi et al. 2022; Pfefferbaum et al. 2023). Normative modeling has been increasingly used in neuropsychiatric imaging analysis and has shown value in highlighting subtle heterogeneity otherwise undetected in classic case-control volumetric comparisons using group-level analyses (Segal et al. 2023; Dima et al. 2022; Tetreault et al. 2020; Rutherford et al. 2023). By employing state-of-the-art harmonization techniques, normative modeling methods aggregate data from multiple databases resulting in models generated from 1000s to tens of 1000s of subjects and covering the human lifespan. To date, these normative models have used cortical thickness or subcortical volumes (Bethlehem et al. 2022; Dima et al. 2022; Potvin et al. 2016; Frangou et al. 2022). To our knowledge, there have not been investigations into the use of normative modeling based on thalamic nuclear volumes (as opposed to whole thalamic volumes). This is in part due to the lack of accurate methods of thalamic nuclei segmentation from structural MRI. Recently, several promising methods for thalamic nuclei segmentation from structural MRI at 3T have been proposed, such as Bayesian estimation from probabilistic atlases (Iglesias et al. 2018), multi-atlas segmentation from specialized white-matter nulled image contrast (Su et al. 2019) or adapted to standard T1 (Bernstein et al. 2021; Vidal et al. 2024). These methods have recently been used to delineate atrophy of *specific* thalamic nuclei in Frontotemporal dementia (Bocchetta et al. 2020; McKenna et al. 2023), Alzheimer’s disease (Low et al. 2019; Bernstein et al. 2021), and alcohol use disorder (Zahr et al. 2020). A very recent work (Williams et al. 2023) which compared these structural methods and evaluated their accuracy found HIPS-THOMAS, a variant of THOMAS adapted for standard T1 MRI to be more accurate and sensitive than other structural MRI based methods.

Empowered by a sensitive and accurate method for analysis of structural T1 MRI data from public databases, we propose the first normative models using thalamic nuclear volumes and examine two disease populations – dementia and psychosis – spanning the younger and older age brackets. The normative models were constructed using control data from 2,336 healthy subjects from multiple public databases spanning the ages of 5 to 100.

## Results

A total of 3,069 of 3097 subjects ages 5.6 to 100 years passed initial QC procedures (Table 1, Figure 1). Of these 3069 subjects, 2,374 belonged to their respective control groups and 695 carried one of the following cognitive or psychiatric diagnoses: early mild cognitive impairment (EMCI), late mild cognitive impairment (LMCI), Alzheimer’s disease (AD), Schizophrenia (SCZ), Bipolar disorder (BD), Attention deficit hyperactivity disorder (ADHD), Affective psychosis (AP), Non-affective psychosis (NAP), Schizophrenia with auditory verbal hallucinations (SCZ +AH), Schizophrenia without auditory verbal hallucinations (SCZ -AH).

**Figure 1.**
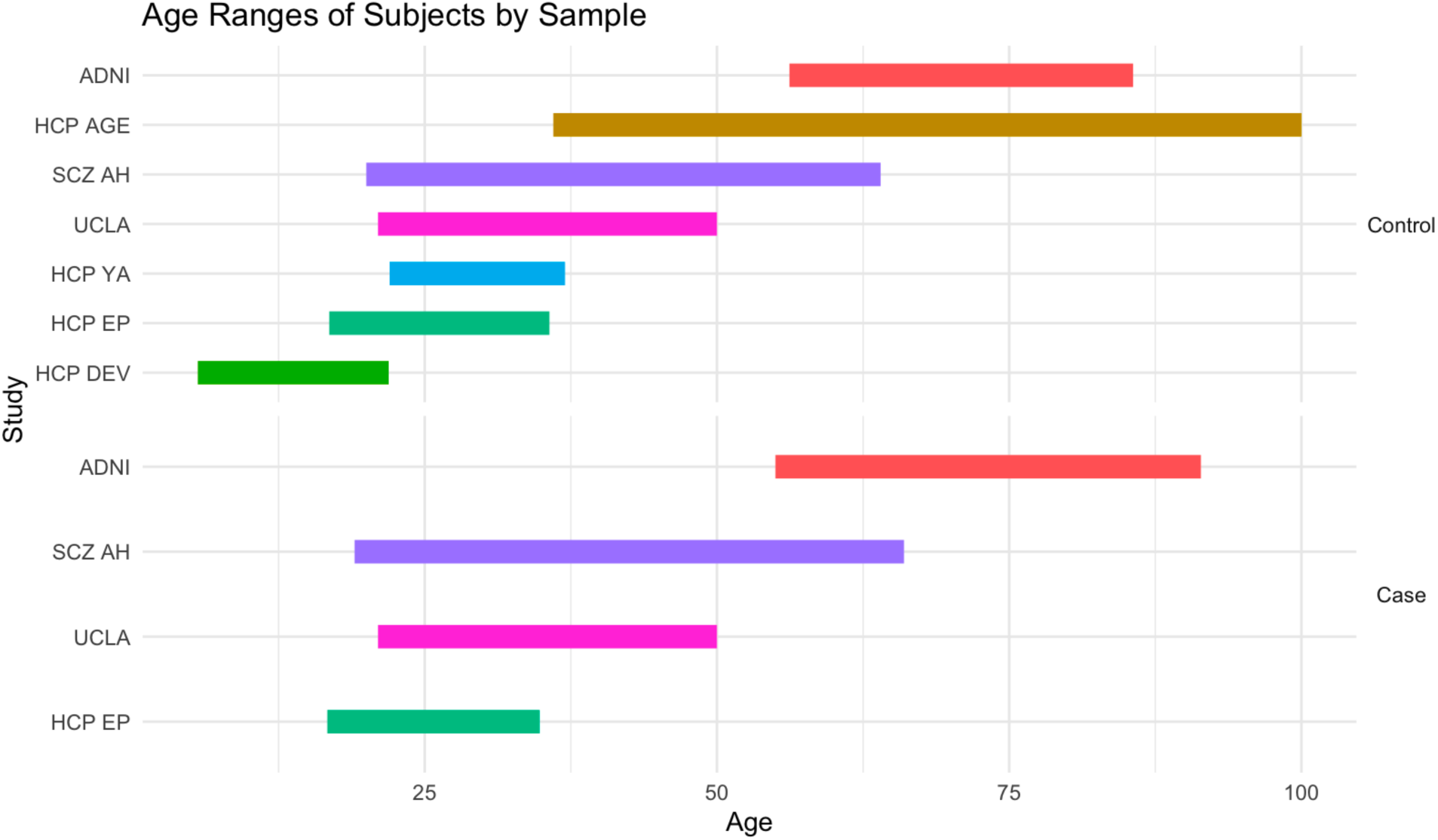
Age ranges for cases and controls within each sample. HCP DEV – The Lifespan Human Connectome Project in Development; HCP EP - The Human Connectome Project for Early Psychosis; HCP YA - Human Connectome Project Young Adult Study; UCLA – UCLA Consortium for Neuropsychiatric Phenomics LA5c Study; SCZ AH - Brain correlates of speech perception in schizophrenia patients with and without auditory hallucinations; SCZ +AH – Schizophrenia with auditory hallucinations; SCZ -AH Schizophrenia without auditory hallucinations; HCP AGE – The Lifespan Human Connectome Project in Aging; ADNI - Alzheimer’s Disease Neuroimaging Initiative; EMCI – Early mild cognitive impairment; LMCI – Late mild cognitive impairment; AD – Alzheimer’s disease.

**Table 1.** Subject characteristics of each sample. HCP DEV – The Lifespan Human Connectome Project in Development; HCP EP - The Human Connectome Project for Early Psychosis; HCP YA - Human Connectome Project Young Adult Study; UCLA – UCLA Consortium for Neuropsychiatric Phenomics LA5c Study; SCZ AH - Brain correlates of speech perception in schizophrenia patients with and without auditory hallucinations; SCZ +AH – Schizophrenia with auditory hallucinations; SCZ -AH Schizophrenia without auditory hallucinations; HCP AGE – The Lifespan Human Connectome Project in Aging; ADNI - Alzheimer’s Disease Neuroimaging Initiative; EMCI – Early mild cognitive impairment; LMCI – Late mild cognitive impairment; AD – Alzheimer’s disease.

| Sample | Phenotype | Mean Age | SD Age | n | Min Age | Max Age | n Male | n Female |
| --- | --- | --- | --- | --- | --- | --- | --- | --- |
| HCP DEV | Control | 14.4 | 4.1 | 628 | 5.6 | 21.9 | 291 | 337 |
| HCP EP | NAP | 22.2 | 3.1 | 83 | 16.7 | 30.8 | 59 | 24 |
|  | AP | 24.5 | 4.3 | 28 | 17.6 | 34.8 | 9 | 19 |
|  | Control | 24.9 | 4.1 | 56 | 16.8 | 35.7 | 37 | 19 |
| HCP YA | Control | 28.7 | 3.7 | 726 | 22.0 | 37.0 | 328 | 398 |
| UCLA | Control | 31.0 | 8.5 | 117 | 21.0 | 50.0 | 62 | 55 |
|  | ADHD | 32.1 | 10.4 | 40 | 21.0 | 50.0 | 21 | 19 |
|  | BD | 35.1 | 8.8 | 46 | 21.0 | 50.0 | 27 | 19 |
|  | SCZ | 36.0 | 9.0 | 41 | 22.0 | 49.0 | 31 | 10 |
| SCZ AH | SCZ +AH | 39.7 | 13.5 | 21 | 19.0 | 66.0 | 19 | 2 |
|  | Control | 41.0 | 14.5 | 22 | 20.0 | 64.0 | 15 | 7 |
|  | SCZ -AH | 45.0 | 7.4 | 23 | 31.0 | 61.0 | 16 | 7 |
| HCP AGE | Control | 60.4 | 15.8 | 701 | 36.0 | 100.0 | 306 | 395 |
| ADNI | EMCI | 70.5 | 7.1 | 211 | 55.5 | 88.6 | 111 | 100 |
|  | LMCI | 71.8 | 7.9 | 114 | 55.0 | 91.4 | 53 | 60 |
|  | Control | 73.4 | 6.2 | 124 | 56.2 | 85.6 | 56 | 68 |
|  | AD | 74.0 | 7.8 | 88 | 55.9 | 88.3 | 47 | 41 |
| Total | Control |  |  | 2374 |  |  |  |  |
|  | Case |  |  | 695 |  |  |  |  |

Six univariate and multivariate models using ordinary least squares regression (OLS), multiple fractional polynomial regression (MFP), and generalized additive models of location, shape, and scale (GAMLSS) were compared with 5-fold cross-validation across the 26 thalamic regions for a total of 780 trained models. Global mean absolute error (MAE) was comparable amongst all models (Table 2). GAMLSS models were the overall top performing models based on MAE. MAE values between test and train datasets were not significantly different as assessed by a two-tailed t-test applied across folds and regions (minimum p=0.93), ruling out overfitting. Training times were 3-4 times longer for multivariate models compared to univariate models and a GAMLSS multivariate model took about 14 seconds to train whereas an MFP model only required 0.2 seconds to train (Supplemental Table 1). Although model performance was similar and GAMLSS requires increased training times, we opted to use the GAMLSS multivariate model as it would increase the public accessibility of our normative models by elimination of steps such as sex-stratification, eTIV adjustment, and combat harmonization and the ability to leverage multiple methods described to adapt a GAMLSS model to new data (Dinga et al. 2021; Bethlehem et al. 2022).

**Table 2.** MAE and rank averaged over all folds, regions, and sexes. MAE was calculated for all models for each fold, region, and sex (male, female, or both for multivariate models). Each model was ranked according to MAE within each fold, region, and sex.

| Model | Mean MAE |  | Mean Rank |  |
| --- | --- | --- | --- | --- |
|  | Train | Test | Train | Test |
| GAMLSS Multivariate | 47.72 | 48.43 | 1.01 | 1.18 |
| GAMLSS Univariate | 47.73 | 48.01 | 1.01 | 1.30 |
| MFP Univariate | 48.10 | 48.45 | 2.04 | 1.95 |
| MFP Multivariate | 48.21 | 49.06 | 1.99 | 2.01 |
| OLS Univariate | 48.84 | 48.98 | 2.95 | 2.75 |
| OLS Multivariate | 49.07 | 49.63 | 3.00 | 2.81 |

Model calibration plots for the GAMLSS multivariate model were visually assessed and did not reveal substantial miscalibration (left Pulvinar shown in Supplemental Figure 2 as an example). Visual inspection of z-scores and volumes demonstrated that the model mitigated effects of age, site, sex, and estimated total intracranial volume (eTIV) as evidenced by near zero slopes for controls and shifting means for cases (Supplemental Figures 3 and 4). Centile plots demonstrated differential developmental and aging trajectories for the thalamic regions. The whole thalamus had a nearly linear decrease in volume proportional to age whereas the lateral geniculate nucleus (LGN) appears to peak around age 30 and the habenula (Hb) has relatively constant volume over the lifespan. Centile curves for the whole thalamus and two example nuclei are shown in Figure 2 (curves for all nuclei are shown in Supplemental Figure 5). Although trajectories over the lifespan differ, z-score distributions for control samples approximate normal distributions for controls for all regions whereas there is a deviation from normal distribution observable in several regions for Alzheimer’s Disease Neuroimaging Initiative (ADNI) data (Figure 3). Additionally, leftward shifts (i.e. atrophy or reduction in volume) in z-score distributions for some regions like anteroventral (AV), medial dorsal-parafascicular (MD-Pf), centromedian (CM), medial geniculate nucleus (MGN), and pulvinar (Pul) can be seen to increase with disease progression from EMCI to LMCI and AD, which likely represents increasing atrophy. Within psychiatric diagnoses, the shifts in z-score distributions were more subtle but there appears to be leftward shifts in the Thalamus and Pul in NAP and the MGN in SCZ +AH (Figure 4, Supplemental Figure 6).

**Figure 2.**
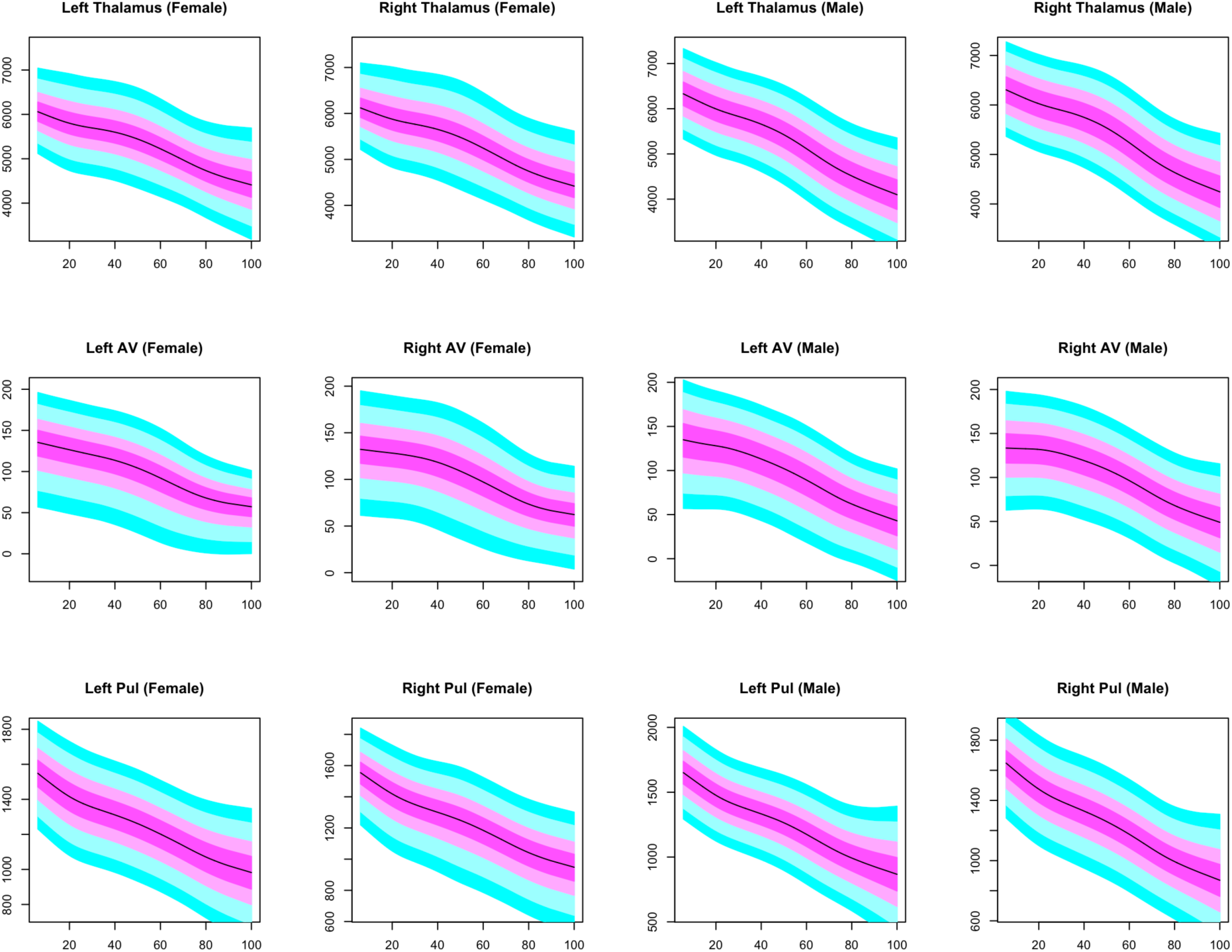
Sex specific centile plots of representative regions for GAMLSS univariate model. Age is on the x-axis, eTIV adjusted volume is on the y-axis.

**Figure 3.**
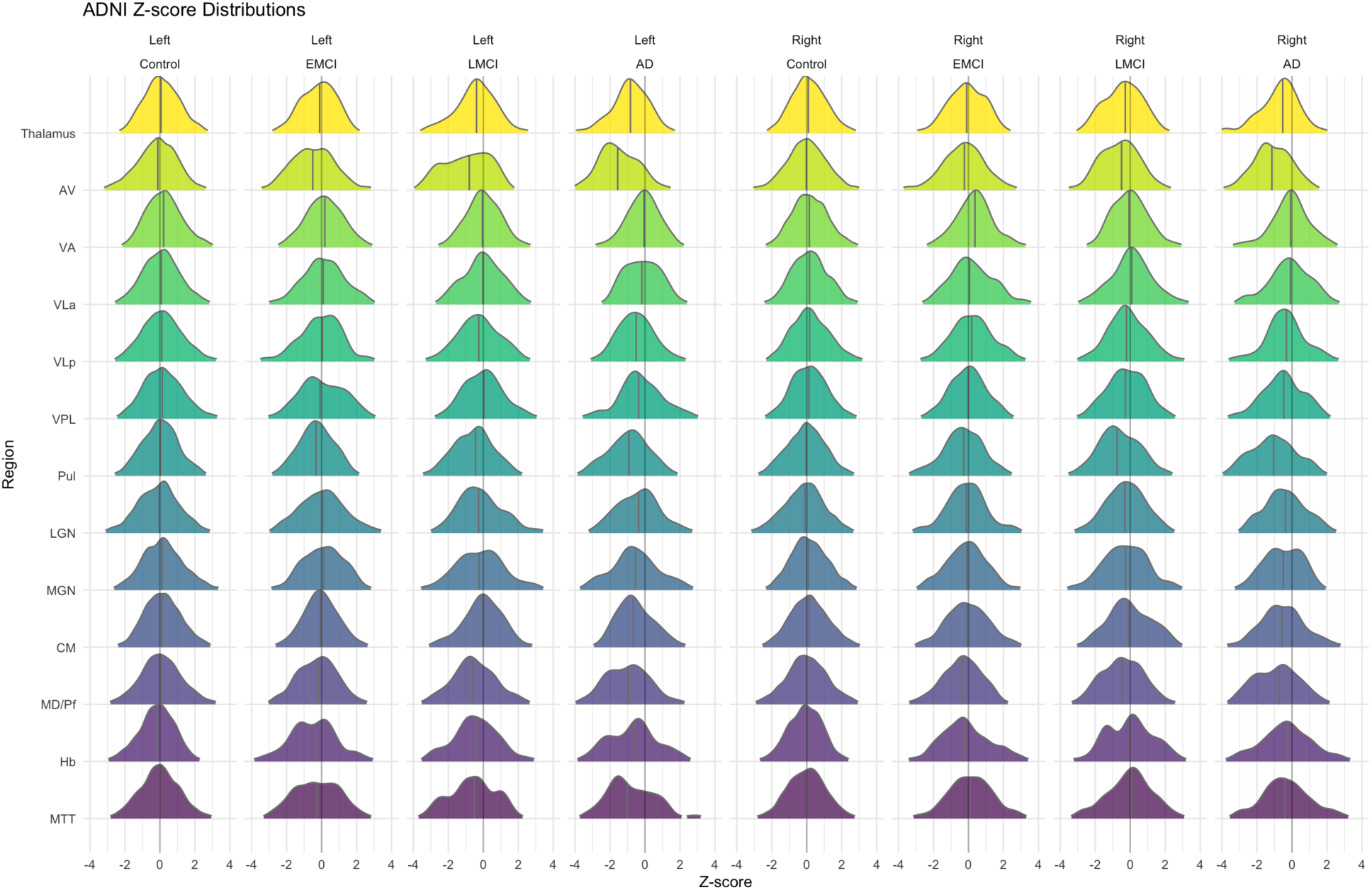
Z-score distributions for the ADNI study. Controls represent the larger pool of control subjects included in the normative model.

**Figure 4.**
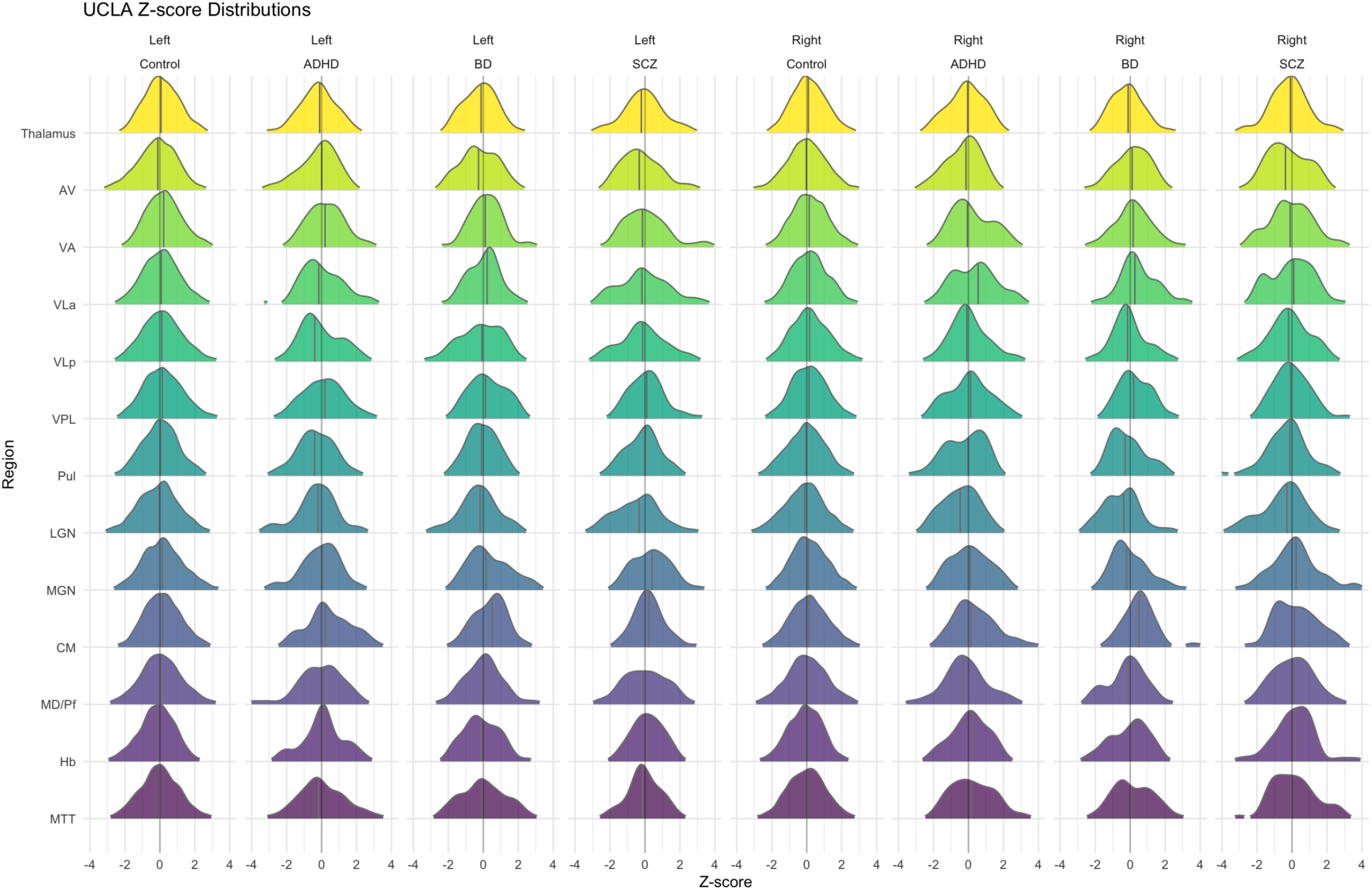
Z-score distributions for the UCLA study including the SCZ, BD, and ADHD phenotypes. Controls represent the larger pool of control subjects included in the normative model.

At least one infranormal z-score and one supranormal z-score were observed in 42.4 percent and 48.4 percent of control subjects respectively (Supplemental Table 2). The percentage of subjects with at least one infranormal z-score increases from EMCI to LMCI to AD while the percentage of individuals with at least one supranormal z-scores decreases. There was a more heterogenous pattern in psychiatric disorders (Supplemental Table 2). In the regional overlap analyses, there was an increase in the percentage of subjects with infranormal z-scores most notably in the AV, Pul, MD-Pf, Hb, and mamilothalamic tract (MTT) and a decrease in the percentage of subjects with supranormal z-scores in most regions that progresses from EMCI to AD (Figure 5). In contrast, psychiatric disorders again demonstrated more heterogeneous patterns of extreme z-score deviations and the regional overlap appears greater for infranormal z-scores than supranormal z-scores for all phenotypes with the possible exception of ADHD and BD (Figure 5).

**Figure 5.**
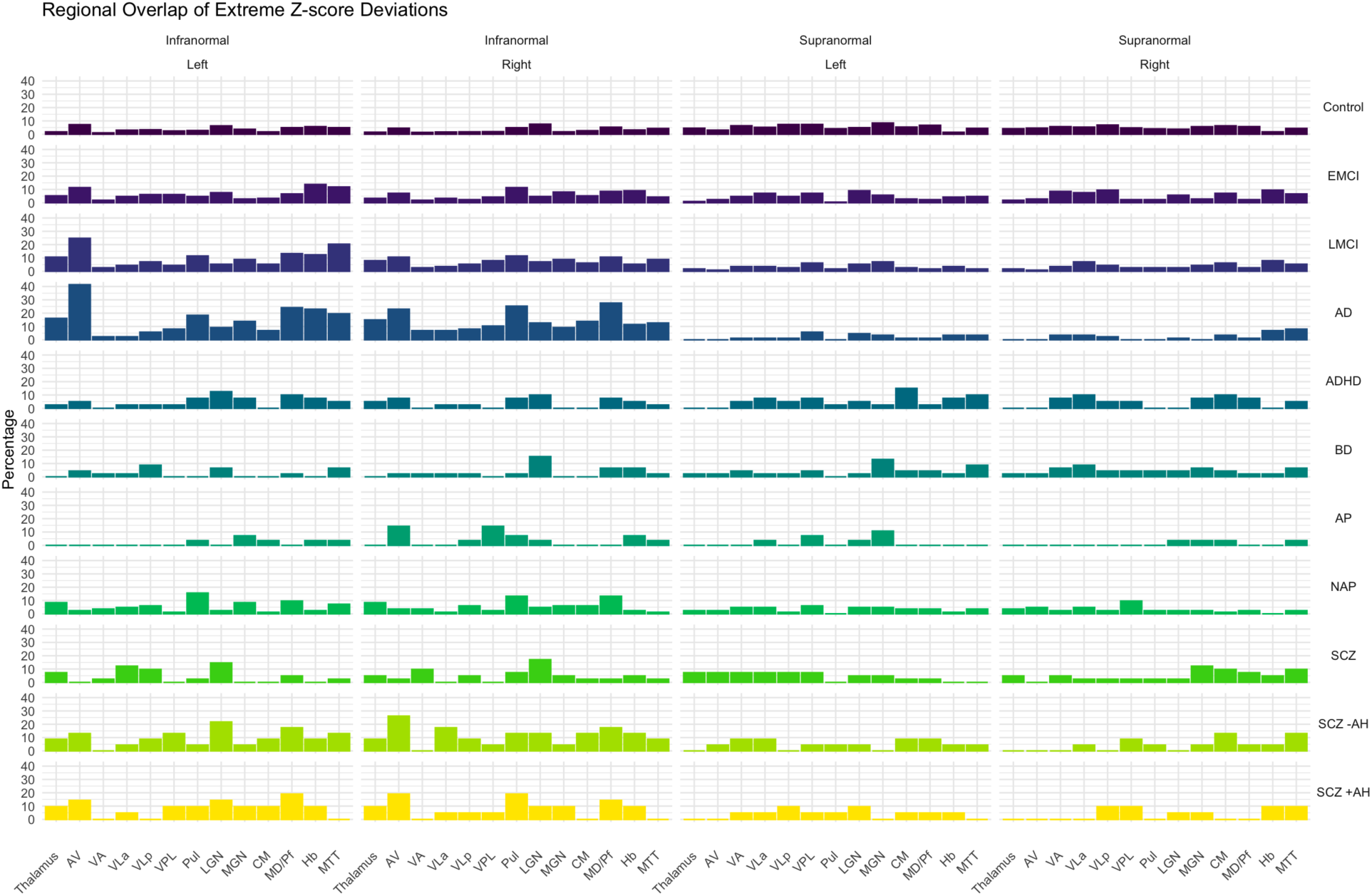
Regional overlap of extreme z-score deviations in cases and controls. Percentage of subjects with extreme z-score deviations in each thalamic region. Infranormal – z-score < -1.96. Supranormal – z-score > 1.96.

In the multigroup comparisons, a statistically significant increase in infranormal z-scores compared to controls was observed in EMCI (p=2.0e-04), LMCI (p=7.3e-09), AD (p<2.22e-16), SCZ -AH (p=4.4e-03), and SCZ +AH (p=9.6e-03) while a statistically significant decrease in supranormal z-scores was observed in AD (p=1.9e-03), AP (p=0.018), NAP (p=0.01) (Figure 6). Analysis of lateralized extreme z-score deviations compared to controls demonstrated a statistically significant increase in infranormal z-scores on the left in EMCI (p=1.5e-04), LMCI (p=3.2e-10), AD (p<2.22e-16), SCZ -AH (p=0.016), and SCZ +AH (p=0.016) and on the right in EMCI (p=4.3e-03), LMCI (p=5.1e-05), AD (p<2.22e-16), SCZ -AH (p=3.1e-03), and SCZ +AH (p=7.7e-03) (Supplemental figure 7). A decrease in supranormal z-score was observed on the left in LMCI (7.7e-03), AD (p=9.3e-04), and NAP (p=0.024) and on the right in AD (p=0.011), AP (p=0.018), and NAP (p=0.048).

**Figure 6.**
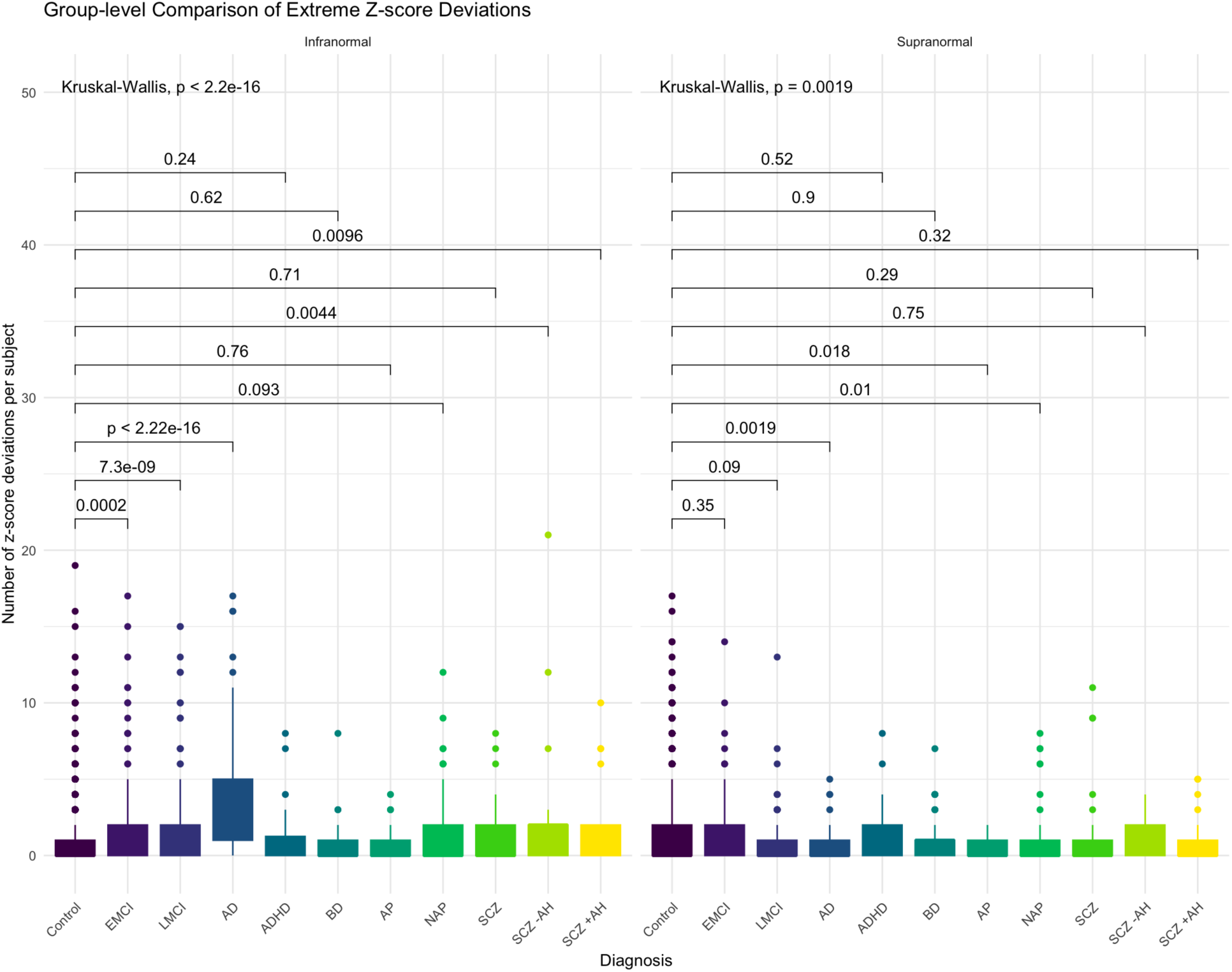
Group-level comparison of extreme z-score deviations between cases and controls.

Statistically significant (FDR adjusted p < 0.05) shifts in z-score distributions were observed in all phenotypes except ADHD and SCZ and all except one shift (CM nulceus in BD) were leftward shifts indicating atrophy (Figure 7). As with the multigroup comparison of lateralized extreme z-score deviations, there is a general trend where the left hemisphere is more affected than the right in EMCI, LMCI, and AD. Specifically, there are significant shifts in EMCI in the left Thalamus, AV, Pul, CM, MD/Pf, Hb, and MTT and in the right Thalamus, AV, Pul, MGN, and MD/Pf. In LMCI, these shifts progress to include the left ventrolateral posterior (VLp) and MGN and right VLp and ventroposterior lateral (VPL); however, the left CM and Hb were not significant. In AD, the shifts increase in magnitude compared to EMCI and LMCI and encompass all regions except the left ventrolateral anterior (VLa) and lateral geniculate nucleus (LGN) and right VLa, LGN, and Hb. All shifts in EMCI, LMCI, and AD had a positive difference between the means of control and case distributions indicating a leftward shift of the distribution consistent with atrophy beyond what would be expected for age.

**Figure 7.**
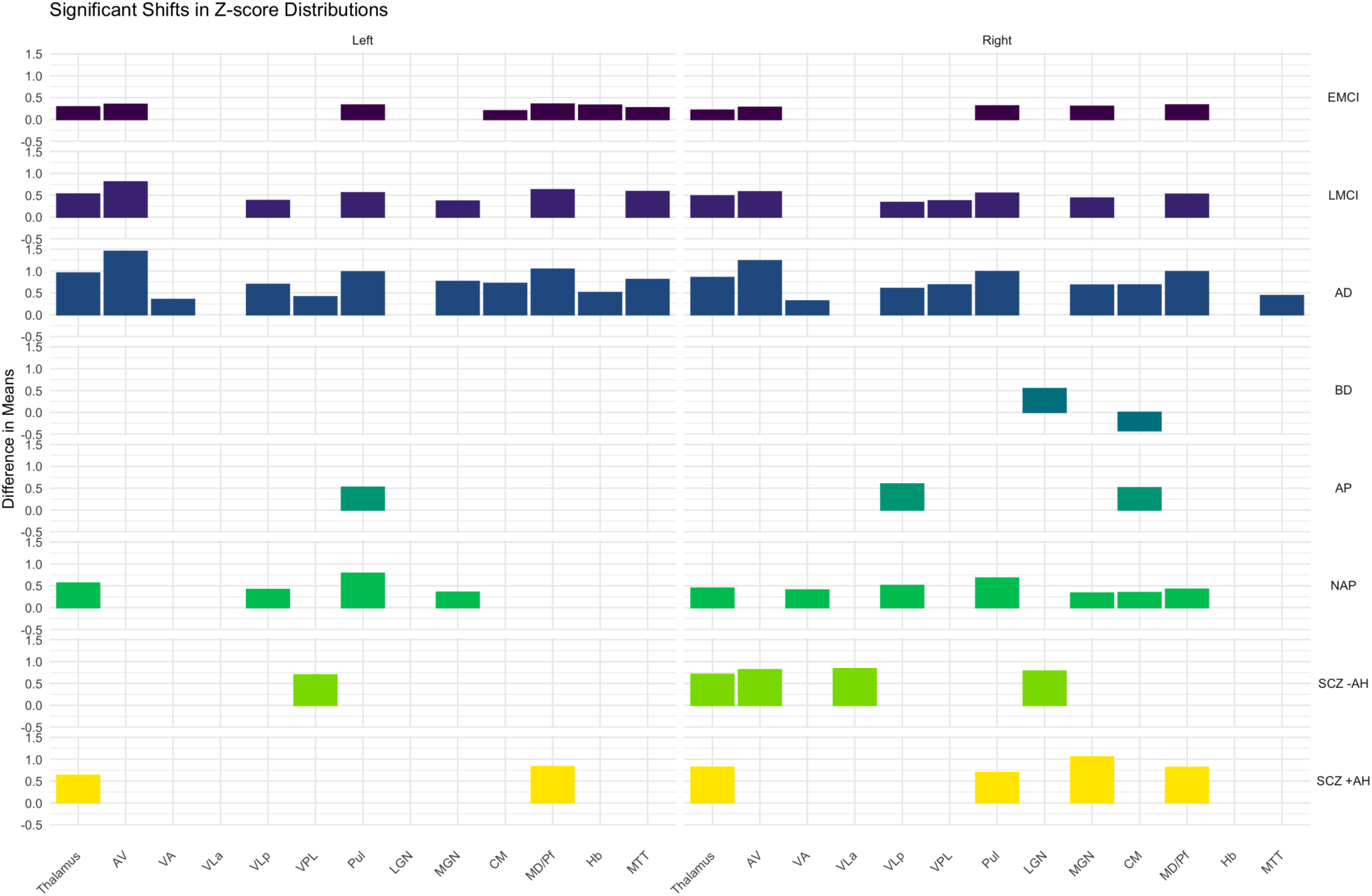
Significant (FDR adjusted p < 0.05) shifts in z-score distributions. The magnitude and direction of the distribution shift is represented as the diderence in the means of z-score distributions between control and cases. There were no significant shifts in ADHD or SCZ from the UCLA study.

In contrast to EMCI, LMCI, and AD, there appears to be a trend where there are more z-score distribution shifts in the right hemisphere compared to the left in psychiatric diagnoses. Within the UCLA Consortium for Neuropsychiatric Phenomics LA5c (UCLA) study (i.e. ADHD, BD, SCZ), there was a significant leftward shift (i.e. atrophy) in the right LGN and a significant rightward shift (i.e. hypertrophy) in the right CM nucleus in BD. The increase in CM was the only right shift (i.e. hypertrophy or increase in volume) observed. There were no shifts in ADHD or SCZ that survived after adjusting for multiple comparisons. Comparing AP and NAP from the Human Connectome Project for Early Psychosis (HCP EP) study, only the left Pul, right VLP, and right CM are shifted in AP while the left Thalamus, VLP, Pul, MGN and right Thalamus, VA, VLp, Pul, MGN, CM, and MD/Pf are shifted in NAP, which suggests the thalamus is more severely affected in NAP compared to AP. Finally, the z-score distribution shifts in SCZ -AH and SCZ +AH are about equal in number and magnitude but have a different pattern. The left VPL and right Thalamus, AV, VLa, and LGN are shift in SCZ -AH while the left Thalamus, left MD/Pf, right Thalamus, right Pul, right MGN, and right MD/Pf are shifted in SCZ +AH.

## Discussion

Normative modelling has shown promise in identifying regional brain abnormalities and delineating subtle heterogeneity often missed in comparisons of mean volumes across groups. Here, we present the development of the first normative model of thalamic nuclear regions, which leverages the flexibility of GAMLSS as a prediction method, the state-of-the-art HIPS-THOMAS for accurate segmentation of thalamic nuclei from standard T1 MRI data, and a large control dataset garnered from 2,366 subjects across 9 cohorts. We also compared the performance of multiple predictive models for normative modeling. Notably, we found that OLS, MFP, and GAMLSS perform similarly. This contrasts with a recent work that found MFP to outperform GAMLSS and OLS in modelling whole subcortical volumes (Ge et al. 2023). This is possibly due to using different modelling strategies, smaller sample size (2,366 in the present study versus 37,407 reported in Ge et al.), or increased variability in Freesurfer-derived subcortical volumes compared to HIPS-THOMAS (Williams et al. 2023). Additionally, we found that the method of controlling for sex, site, and eTIV had minimal impact on model performance, which has not been previously investigated. While GAMLSS was the best performing model by a small margin, utilizing a multivariate model where the random effects of site can be estimated for new data without retraining the model will simplify data preparation, potentially enhance useability by the general public, and preserves a larger sample size by avoiding sex-stratification. It should also be noted that we observed that GAMLSS can get stuck in local minima, which can lead to failure of convergence and increased training times. Models using Combat also incurred additional computational time.

In addition to the optimization of our normative model, our analysis of the ADNI cohort (i.e. EMCI, LMCI, and AD) supports the validity of our models. Although there are several phenotypes of AD, an estimated 78% of AD patients have a typical amnestic syndrome (Dubois et al. 2023). Additionally, up to 13% of patients with MCI progress to AD annually (McGirr et al. 2022). In our regional overlap analyses, group level analyses of extreme z-score deviations, and analysis of shifts in z-score distributions, we saw a clear progression of decreasing z-scores from EMCI to LMCI and AD including AV, VA, pulvinar, CM, MD, MGN, and MTT. These results are congruent with prior ANCOVA-based analyses using the same ADNI data by Bernstein et al. 2021. They also comport with the typical progression of amyloid and tau from the medial temporal lobes to the medial prefrontal and parietal lobes, and observations of both subcortical spread of amyloid and deposition of amyloid and tau in the anterior thalamic nuclei (Bernstein et al. 2021; Busche and Hyman 2020; Cho et al. 2018; Aggleton et al. 2016). Interestingly, there is evidence of alterations in the MGN prior to the development of neurofibrillary tangles and senile plaques (Bartlett 2013). We also observed that left thalamic regions were more affected than the right consistent with evidence that grey matter atrophies faster and earlier in the disease on the left (Lubben et al. 2021). Taken together this is compelling evidence that our normative model is well calibrated and captures volumetric changes associated with underlying pathology and disease state.

In contrast to dementia and consistent with other efforts (Segal et al. 2023; Worker et al. 2023; Antoniades et al. 2021; Lv et al. 2021), we found significant heterogeneity in psychiatric disorders. Regional overlap analyses, group level analyses of extreme z-score deviations, and analysis of shifts in z-score distributions, which include individuals carrying a diagnosis of a schizophrenia spectrum disorder (NAP, SCZ, SCZ -AH, SCZ +AH) in three separate samples all showed heterogeneity.

There are some notable results for specific diagnoses including BD and SCZ +/- AH. In BD, there was a leftward shift (atrophy) in the right LGN and a rightward shift (hypertrophy) in the right CM. The LGN is a sensory-specific nucleus that primarily relays visual information and receives input from the retina and the primary visual cortex. However, it also receives input from cholinergic, serotonergic, and noradrenergic centers and may also be involved in the modulation of circadian rhythms via projections to the suprachiasmatic nucleus and pineal gland (Covington and Al Khalili 2023). The CM plays a major role in attention, arousal, and sensorimotor learning through its connections to subcortical structures including the basal ganglia; the premotor, motor, and somatosensory cortices; the brainstem; and other thalamic nuclei (Ilyas et al. 2019). All subjects in the BD group carried a diagnosis of Bipolar I Disorder indicating that all subjects had experienced at least 1 manic episode, which includes the symptoms of an increased level of alertness and decreased need for sleep (American Psychiatric Association, Association, and Others 2013). Here, the role of the LGN and CM in arousal and circadian rhythms suggest a potential role in mania with hypertrophy of the CM suggesting overactivity and/or increased connectivity.

Group level analysis of SCZ -AH and SCZ +AH demonstrated a statistically significant increase in infranormal z-scores in both groups as compared to controls. Within the lateralized group comparison, the p-values are more significant for both SCZ -AH and SCZ +AH on the right (Supplemental Figure 7). In the SCZ +AH group, the MGN and Pul had a leftward shift on the right side and MD demonstrated bilateral leftward shifts, which are not present in the SCZ -AH group (Figure 7). In contrast, SCZ-AH showed shifts in the left VPL, right AV, right VLa, and right LGN. Like the LGN, the MGN is a small, sensory-specific nuclei that is primarily associated with transmitting auditory information from the brainstem to the primary auditory cortex. However, it has reciprocal connections with both the primary and secondary auditory cortices, projects to the amygdala, and actively modulates and filters incoming auditory information (Bartlett 2013). In contrast, Pulvinar (Pul) is a large higher-order associative nucleus with extensive connections to the prefrontal cortices, posterior parietal cortex, occipital cortex, amygdala, and heteromodal association cortices suggesting that the Pul coordinates activity in distributed networks including the thalamocortical auditory circuit (Benarroch 2015; Barczak et al. 2018). Similarly, MD is a higher-order association nucleus with reciprocal connections to the prefrontal cortex, basal ganglia, and amygdala that can relay multisensory information to the prefrontal cortex and contribute to the coordination of thoughts, behavior and language (Georgescu, Popa, and Zagrean 2020; Schmahmann 2003; Barbas, García-Cabezas, and Zikopoulos 2013). While there are many types of auditory hallucinations, all the SCZ +AH subjects experienced auditory verbal hallucinations (AVH) while the SCZ -AH subjects did not. AVH are often subjectively experienced as being from an external or an internal source; can take the form of commands, comments, or narrations; are often emotionally charged; may represent subconscious thought or subvocalizations misattributed to an external agent; and are potentially mediated by abnormalities in a large scale auditory network that includes the thalamus, auditory cortex, and the language centers (Blom 2015; Thakkar, Mathalon, and Ford 2021). These results suggest that volumetric changes in the thalamus are associated with subtle abnormalities in the thalamus-regulated networks that integrate and mediate the perception of sound, movement, motivated behavior, language, and emotion, which accounts for diversity in the subjective experience of AVH.

Both AP and NAP showed decreases in supranormal z-scores that have a slight preference for the right in AP and the left in NAP based on smaller p-values. Both groups also show shifts in z-score distributions in the left Pul, right VLP, and right CM while NAP has additional shifts in the left Thalamus, VLP, and MGN and right Thalamus, VA, Pul, MGN, and MD/Pf, which suggests that NAP has greater pathological changes in the thalamus as compared to AP and that the right thalamus is more affected than the left. These results highlight the difference between identifying extreme z-score deviations versus a shift in the overall z-score distribution in disease states. As previously discussed, there are multiple mechanisms by which the Pul, MGN, CM, and MD may lead to psychosis, particularly AVH. The diagnostic criteria for primary psychotic disorders, however, spans a range of symptoms including delusions, hallucinations, disorganization (thought, language, behavior), and negative symptoms (blunted affect, avolition, anhedonia, asociality, alogia), which similarly implicate frontal-subcortical circuits involving the VA and VLP (Strauss and Cohen 2017; Bonelli and Cummings 2007; Vandevelde et al. 2018).

Regional overlap of extreme z-score deviations (Figure 5) suggests the identical nuclei may engage in different disease states, which aligns with the earlier observations concerning the functional and behavioral multiplicity nature of relay and higher-order thalamus nuclei (Kumar et al. 2022). Interestingly, these highly deviated nuclei belong to relay and higher-order thalamus nuclei, hinting at an impact of volume alteration in both thalamocortical and cortico-cortical functional processing via thalamic nuclei.

These findings are consistent with recent volumetric analyses of thalamic nuclear regions comparing schizophrenia and bipolar disorder and schizophrenia with and without hallucinations to healthy controls, which identified significant associations with the MD, Pul, LGN, and MGN (Mørch-Johnsen et al. 2023; Perez-Rando et al. 2022). In addition, we have identified volumetric reductions affecting the VA, VLP, and CM. This is potentially due to difference in the method used to segment the thalamus and comparison to a larger control population. There also appears to be a slight right hemispheric predilection in z-score distribution shifts across all the psychiatric diagnoses. Functional and structural asymmetry of the brain are well described and it is likely that lateralized findings represent differences in underlying psychopathology (Lubben et al. 2021; Low et al. 2019; Tomer et al. 2013; Tiihonen et al. 1998; Kuo and Massoud 2022).

The strengths of our normative modeling-based analyses include the use of the robust state-of-the-art methodology for both thalamic segmentation and normative modelling, a large control sample size, optimization and validation of the normative model using well-defined cognitive disorders, significant results consistent with prior analyses in psychiatric diagnoses, and volumetric changes in the thalamus that potentially explain psychopathology based on known functional neuroanatomy. Importantly, these insights could not be made based on just volumetric analysis of the whole thalamus as has been typically done. Additionally, normative modelling has successfully addressed the effects of individual variation like age, sex, eTIV, and site simplifying the interpretation in terms of disease state. As the role of the thalamus in MCI, AD, and psychiatric disease remains an area of active research, increasing the use of HIPS-THOMAS in conjunction with normative modelling can serve a critical role in future research, particularly if extended beyond volumetric analyses to include structural and/or functional connectivity measures.

Weaknesses include a smaller number of individuals for some psychiatric diagnoses, fewer control samples in the older age ranges, less aggressive removal of outliers from the control groups, absence of some thalamic nuclei like centrolateral or intralaminar nuclei in the analyses, and using the same model for all regions. While +/- 1.5 IQR is a typical threshold for outlier removal, we wanted to use a more conservative range to avoid bias in the group level analysis and z-scores. This potentially affected model calibration, however, our validation procedure and results that are consistent with those previously reported suggest this did not have a major impact. Additionally, our analyses of the MCI and AD continuum suggest that our model performed well even in the older age ranges. Several thalamic nuclei including MD and Pul have been delineated into subregions with differential functions and connectivity. This would require integration of HIPS-THOMAS with diffusion or resting state fMRI based parcellation which is challenging and remains an area for future work. Our interpretation of volumetric changes is dependent on a change in volume being associated with a change in network function without assessing the network directly. While this is another area of future work, the concept of Hebbian learning, the diaschisis hypothesis, and emerging evidence that atrophy propagates along network lines support this approach (Munakata and Pfaffly 2004; Carrera and Tononi 2014; Han et al. 2023; Petersen et al. 2022; Chopra et al. 2023). Some regions like the LGN, MGN, CM, Hb, and MTT are small and potentially more susceptible to subtle variability and imaging artifacts despite their accurate segmentation. Results concerning these nuclei have to be interpreted with caution.

There are many sources of heterogeneity in psychiatric disease including changes in diagnostic criteria over time, variability in how diagnostic criteria are applied, and comorbidity with other psychiatric diagnoses including trauma and substance use (Tandon et al. 2013; Regier et al. 2013; Russell et al. 2024; Martinotti, Fornaro, and De Berardis 2023). This heterogeneity extends even to a relatively focused symptom like AVH (Blom 2015; Thakkar, Mathalon, and Ford 2021). Normative modelling is a potential means of addressing heterogeneity as it allows direct comparison between individuals and between distinct regions within individuals, however, its application has been limited to group-level analysis. Future work is needed to address the individual and more specific transdiagnostic psychopathology. One challenge will be in identifying z-scores that are abnormal for the individual but do not meet the criteria for an extreme z-score deviation. Moreover, a significant percentage of control samples were found to have extreme z-score deviations, which have unclear significance. Normative modelling of the thalamus can also be extended to additional subcortical structures, connectivity, asymmetry, and association with clinical measures.

## Conclusion

Here we present the first normative model of thalamic nuclear volumes along with analyses to support its validity, consistency with other volumetric analyses, and potential to generate novel insight into disease states. We have also undertaken one of the few comparisons of normative modeling methods and the only comparison to-date of univariate and multivariate normative models, which demonstrates interchangeability of the two. Additionally, our centile curves provide important insight into volume changes to thalamic nuclear regions across the lifespan, where there is a paucity of research. Models and code will be available at https://github.com/thalamicseg/hipsthomasdocker

## Methods

### Imaging Data Acquisition and Sample Populations

Unprocessed T1w images and clinical measures were accessed from the following resources:

### The Lifespan Human Connectome Project in Development (HCP DEV)

All subjects were defined as controls and considered to have a broadly typical development that excluded a history of a serious medical or psychiatric condition (Harms et al. 2018; Somerville et al. 2018).

### Human Connectome Project Young Adult Study S900 Release (HCP YA)

All subjects belong to a sibship, were defined as controls, and considered to be broadly healthy without a history of neurodevelopmental, psychiatric, or neurological disorders (Van Essen et al. 2012).

### The Lifespan Human Connectome Project in Aging (HCP AGE)

All subjects were defined as controls and had typical aging without major psychiatric or neurologic disorders. Subjects were screened for cognitive deficits and while dementia like Alzheimer’s disease was excluded, mild to moderate cognitive deficits were allowed depending on the age of the subject (Bookheimer et al. 2019).

### The Human Connectome Project for Early Psychosis (HCP EP)

All subjects were assessed with the SCID-5-RV. Control subjects did not have formal psychiatric diagnoses or history of psychiatric hospitalization. Subjects with Non-affective psychosis (NAP) met DSM V criteria for schizophrenia, schizophreniform, schizoaffective, psychosis NOS, delusional disorder, or brief psychotic disorder with onset within five years of study enrollment. Subjects with Affective psychosis (AP) met DSM V criteria for major depression with psychosis or bipolar disorder with psychosis with onset within five years of study enrollment (Lewandowski et al. 2020).

### UCLA Consortium for Neuropsychiatric Phenomics LA5c Study (UCLA)

Control subjects were without prior psychiatric diagnosis including substance abuse. Case subjects were diagnosed with ADHD, bipolar disorder, or schizophrenia according to criteria in the DSM IV using the SCID-I. OpenNeuro Dataset ds000030 (Bilder et al. 2020; Poldrack et al. 2016).

### Brain Correlates of Speech Perception in Schizophrenia Patients With and Without Auditory Hallucinations (SCZ AH)

All subjects were assessed with the SCID-5-RV. Control subjects were age and gender matched to cases without past psychiatric diagnoses. Cases were defined as those meeting DSM V criteria for schizophrenia or schizoaffective disorder and either experienced daily auditory verbal hallucinations (SCZ +AH) or were free of hallucinations for at least 6 months (SCZ -AH). OpenNeuro Dataset ds004302 (Soler-Vidal et al. 2022).

### The Alzheimer’s Disease Neuroimaging Initiative (ADNI)

Controls were cognitively normal subjects without memory concerns, evidence of dementia, MMSE >= 24, and education adjusted Wechsler Memory Scale Logical Memory II ≥ 9 for 16 or more years of education, ≥5 for 8-15 years of education, ≥3 for 0-7 years of education. Early Mild Cognitive Impairment (EMCI) was defined as having subjective memory complaint without functional impairment and MMSE >= 24. The status of “early” MCI was established using education adjusted scores on the Wechsler Memory Scale Logical Memory II ≥16 years education: 9-11; 8-15 years: 5-9; 0-7 years: 3-6. Late Mild Cognitive Impairment (LMCI) was defined as having subjective memory complaint without functional impairment and MMSE >= 24. The status of “late” MCI was established using the Wechsler Memory Scale Logical Memory II ≥16 years of education: ≤8; 8-15 years: ≤4; 0-7 years: ≤2. Alzheimer’s Disease (AD) met definition of probable Alzheimer’s disease based on National Institute of Neurological and Communicative Disorders and Stroke (NINCDS) / Alzheimer’s Disease and Related Disorders Association (ADRDA) criteria and MMSE 20-26 (Beckett et al. 2015). A subset of 540 subjects acquired on Siemens 3T were used as described in Bernstein et al.

### Thalamic nuclei segmentation

Thalamus-optimized multi-atlas segmentation (THOMAS) is a state-of-the-art method proposed for accurate segmentation of thalamic nuclei, leveraging the superior intrathalamic contrast of white-matter nulled (WMn) MPRAGE data. To segment standard T1 MPRAGE data which is the structural MRI sequence used in most clinical protocols and in public databases, a variant was recently proposed that uses a Histogram-based polynomial synthesis (HIPS) to first generate WMn-like images from T1 prior to segmentation. This variant termed HIPS-THOMAS was shown to significantly improve accuracy characterized using Dice and volume similarity indices (Vidal et al. 2024). Briefly, input T1 images are N4-bias corrected to remove shading artifacts and then automatically cropped to extract a 3D volume encompassing both thalami and then converted to WMn-like contrast using a polynomial transformation. This synthesized image is then segmented using the multi-atlas pipeline of THOMAS, leveraging the improved intrathalamic contrast. THOMAS uses 20 manually labelled high-resolution WMn-MPRAGE datasets (0.8x0.8x1 mm acquired at 7T) which are transferred to the input data space via a high-resolution template space using diffeomorphic nonlinear registration. A joint label fusion algorithm is used to fuse the 20 sets of labels to generate the final thalamic nuclei segmentation. This is summarized in Supplementary Figure 1. Ten thalamic nuclei, the whole thalamus, the habenula (Hb), and the mammillothalamic tract (MTT) volumes were generated for each hemisphere. The ten nuclei can be grouped per region as follows:

1. Anterior group: Antero-ventral (AV)
2. Posterior group: Pulvinar (Pul), Lateral and medial geniculate nuclei (LGN, MGN)
3. Medial group: Mediodorsal-parafasicular (MD/Pf)
4. Intralaminar: Centromedian (CM)
5. Lateral group: Ventral Anterior (VA), Ventral lateral Anterior (VLa), Ventral lateral Posterior (VLp) and Ventral posterolateral (VPL)

### Data quality control

All segmentations were visually inspected using a custom Python script that generated a montage of axial, sagittal, and coronal slices of the thalamic nuclear segmentations overlaid on cropped input images for rapid review. Subjects with missing or failed segmentations were reprocessed or excluded.

### Data preprocessing

Volumes for control samples +/- 3X the interquartile range (IQR) for each site were retained. This larger threshold was chosen to establish a more conservative estimate (compared to the conventional 1.5X IQR) of model performance and subsequent z-scores. To generate data for univariate models (see below), the following preprocessing was performed: volumes were first corrected for total intracranial volume (TIV), site effects were mitigated with Combat in sex-stratified data sets as implemented in NeuroHarmonize python library (Johnson, Li, and Rabinovic 2007; Fortin et al. 2018; Pomponio et al. 2020), volumes were then normalized by subtracting the mean (mean centering), and finally sex-stratified datasets were created. For TIV correction, the estimated total intracranial volume (eTIV) was generated using the Freesurfer *mri_segstats* binary (Buckner et al. 2004). Volumes were adjusted for eTIV using the residual method (Dima et al. 2022; Mathalon et al. 1993) as follows:

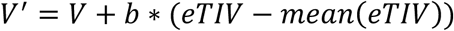

Here V’ and V are the adjusted and non-adjusted volumes and *b* is the slope of V vs. eTIV estimated as the covariance(eTIV,V)/variance(eTIV). Volumes were adjusted separately for each region and site. The above preprocessing steps were omitted for multivariate models and sex, eTIV, and site were used as covariates.

### Normative Modeling Methods and Optimization

Very few comparisons of predictive models for normative modeling have been reported (Ge at al.). We compared 3 different predictive models – ordinary least squares regression (OLS), multiple fractional polynomial regression (MFP), and generalized additive models of location shape and scale (GAMLSS) (Bethlehem et al. 2022; Dima et al. 2022; Dinga et al. 2021; Bozek et al. 2023) which have been previously described well for normative modeling. For each method, we tested a univariate and a multivariate model. In the univariate models, age was the predictive variable, and the models were trained on sex stratified data sets following eTIV volume adjustment, site harmonization with Combat, and mean-centering as described above. A 5-fold cross-validation was used to evaluate each model. All models were implemented in R and trained for each region independently. The 3 models are briefly described in the supplemental text.

The model with the best overall performance based on average mean absolute error (MAE) was selected and trained on all control samples for each region. Z-scores were calculated for cases and controls from the residuals of fitted values and the standard deviation of residuals in the control samples. In the case of GAMLSS, fitted values were predicted for both μ and σ for each observation. Z-scores were calculated as follows:

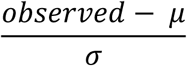

Extreme z-score deviations (infranormal and supranormal) were calculated as those outside the 95^th^ percentile (|z| > 1.96). Regional overlap was calculated for each phenotype as the percentage of subjects within each phenotype with an extreme z-score deviation in a given region. Multigroup comparison between cases and controls was performed using the Kruskal-Wallis rank sum test on counts of extreme z-score deviations. Z-score distributions between cases and controls were compared with the z-test for each region. To control for multiple hypothesis testing, p-values were adjusted using the Benjamini and Hochberg method, and a false discovery rate (FDR) threshold of FDR adjusted p<0.05 was set.

## Data Availability

The authors will attempt to make all data available upon reasonable request. There may be limitations on the distribution of downstream products from the Human Connectome Project and Alzheimer's Disease Neuroimaging Initiative.

https://github.com/thalamicseg/hipsthomasdocker

https://www.humanconnectome.org

https://adni.loni.usc.edu

https://openneuro.org/datasets/ds000030/versions/1.0.0

https://openneuro.org/datasets/ds004302/versions/1.0.1

## Acknowledgements

Data were provided [in part] by the Human Connectome Project, WU-Minn Consortium (Principal Investigators: David Van Essen and Kamil Ugurbil; 1U54MH091657) funded by the 16 NIH Institutes and Centers that support the NIH Blueprint for Neuroscience Research; and by the McDonnell Center for Systems Neuroscience at Washington University.

Research reported in this publication was supported by the National Institute Of Mental Health of the National Institutes of Health under Award Number U01MH109589 and by funds provided by the McDonnell Center for Systems Neuroscience at Washington University in St. Louis. The HCP-Development 2.0 Release data used in this report came from DOI: 10.15154/1520708.

Research reported in this publication was supported by the National Institute On Aging of the National Institutes of Health under Award Number U01AG052564 and by funds provided by the McDonnell Center for Systems Neuroscience at Washington University in St. Louis. The HCP-Aging 2.0 Release data used in this report came from DOI: 10.15154/1520707.

Research using Human Connectome Project for Early Psychosis (HCP-EP) data reported in this publication was supported by the National Institute of Mental Health of the National Institutes of Health under Award Number U01MH109977. The HCP-EP 1.1 Release data used in this report came from DOI: 10.15154/1522899.

*Data used in preparation of this article were obtained from the Alzheimer’s Disease Neuroimaging Initiative (ADNI) database (adni.loni.usc.edu). As such, the investigators within the ADNI contributed to the design and implementation of ADNI and/or provided data but did not participate in analysis or writing of this report. A complete listing of ADNI investigators can be found at: http://adni.loni.usc.edu/wp-content/uploads/how_to_apply/ADNI_Acknowledgement_List.pdf

Data collection and sharing for this project was funded by the Alzheimer’s Disease Neuroimaging Initiative (ADNI) (National Institutes of Health Grant U01 AG024904) and DOD ADNI (Department of Defense award number W81XWH-12-2-0012). ADNI is funded by the National Institute on Aging, the National Institute of Biomedical Imaging and Bioengineering, and through generous contributions from the following: AbbVie, Alzheimer’s Association; Alzheimer’s Drug Discovery Foundation; Araclon Biotech; BioClinica, Inc.; Biogen; Bristol-Myers Squibb Company; CereSpir, Inc.; Cogstate; Eisai Inc.; Elan Pharmaceuticals, Inc.; Eli Lilly and Company; EuroImmun; F. Hoffmann-La Roche Ltd and its affiliated company Genentech, Inc.; Fujirebio; GE Healthcare; IXICO Ltd.;Janssen Alzheimer Immunotherapy Research & Development, LLC.; Johnson & Johnson Pharmaceutical Research & Development LLC.; Lumosity; Lundbeck; Merck & Co., Inc.;Meso Scale Diagnostics, LLC.; NeuroRxResearch; Neurotrack Technologies; Novartis Pharmaceuticals Corporation; Pfizer Inc.; Piramal Imaging; Servier; Takeda Pharmaceutical Company; and Transition Therapeutics. The Canadian Institutes of Health Research is providing funds to support ADNI clinical sites in Canada. Private sector contributions are facilitated by the Foundation for the National Institutes of Health (www.fnih.org). The grantee organization is the Northern California Institute for Research and Education, and the study is coordinated by the Alzheimer’s Therapeutic Research Institute at the University of Southern California. ADNI data are disseminated by the Laboratory for Neuro Imaging at the University of Southern California.

Research using Human Connectome Project for Early Psychosis (HCP-EP) data reported in this publication was supported by the National Institute of Mental Health of the National Institutes of Health under Award Number U01MH109977. The HCP-EP 1.1 Release data used in this report came from DOI: 10.15154/1522899.

All authors have reviewed and approved this manuscript.

