## Supplemental Methods, Figures, and Tables for "Normative modeling of thalamic nuclear volumes"

#### Normative Modeling

Ordinary least squares regression (OLS) was implemented using the *lm* function in base R and takes the following forms for univariate and multivariate models:

$$V = \beta_0 + \beta_1 \cdot age + \varepsilon$$

$$V = \beta_0 + \beta_1 \cdot age + \beta_2 \cdot sex + \beta_3 \cdot eTIV + \beta_4 \cdot site + \varepsilon$$

Multiple fractional polynomial regression (MFP) was implemented using the *mfp* and *fp* functions from the *mfp* package. Briefly, *fp* defines a power expansion of a given variable using powers from the set S = {-2, -1, -0.5, 0, 0.5, 1, 2, 3} and the specified degrees of freedom defining the number of power combinations:

$$f_p(X) = X^{p1} + X^{p2} + \cdots + X^{pi}$$

Where $X$ is a continuous variable, $p$ is a power from set S, and $i$ is the specified degrees of freedom. *mfp* uses an iterative backward selection method to keep only significant variables and powers. Here, we used the maximum degrees of freedom *i*=4. Univariate and multivariate models were as follows:

$$V = \beta_0 + \beta_1 \cdot f_p(age) + \varepsilon$$

$$V = \beta_0 + \beta_1 \cdot f_p(age * female) + \beta_2 \cdot f_p(age * male) + \beta_3 \cdot f_p(eTIV * female) + \beta_4 \cdot f_p(eTIV * male) + \beta_5 \cdot site + \varepsilon$$

Generalized additive models of location shape and scale (GAMLSS) was implemented using the *gamlss, cs,* and *random* functions from the *gamlss* package in R. Briefly, *gamlss* models the mean ($\mu$), standard deviation ($\sigma$), skew ($\nu$), and curtosis ($\tau$) over a distribution, *cs* fits a cubic smoothing spline, and *random* fits random effects. The GAMLSS models take the following forms:

$$V \sim SHASH(\mu, \sigma, \nu, \tau)$$
$$\mu_u = \beta_0 + f_{cs}(age)$$
$$\mu_m = \beta_0 + f_{cs}(age * female) + f_{cs}(age * male) + f_{cs}(eTIV * female) + f_{cs}(eTIV * male) + Zsite$$
$$\sigma = \beta_0 + f_{cs}(age)$$
$$\nu = \beta_\upsilon$$
$$\tau = \beta_\tau$$

Where $SHASH$ is the sinh-arcsinh distribution, $\mu_u$ is a univariate model, $\mu_m$ is a multivariate model, $f_{cs}$ is a cubic smoothing spline, and $Z$ are random effects intercepts.

**Model Training Time**

The *microbenchmark* R package was used to evaluate training times for all models. Each model was trained 20 times on the control samples using volumes for the left whole thalamus. Training times were then averaged. The hardware was an Apple MacBook Pro with Apple M1 Max processor (8 performance cores, 2 efficiency cores) with 64 GB of ram.

### Supplemental Figures

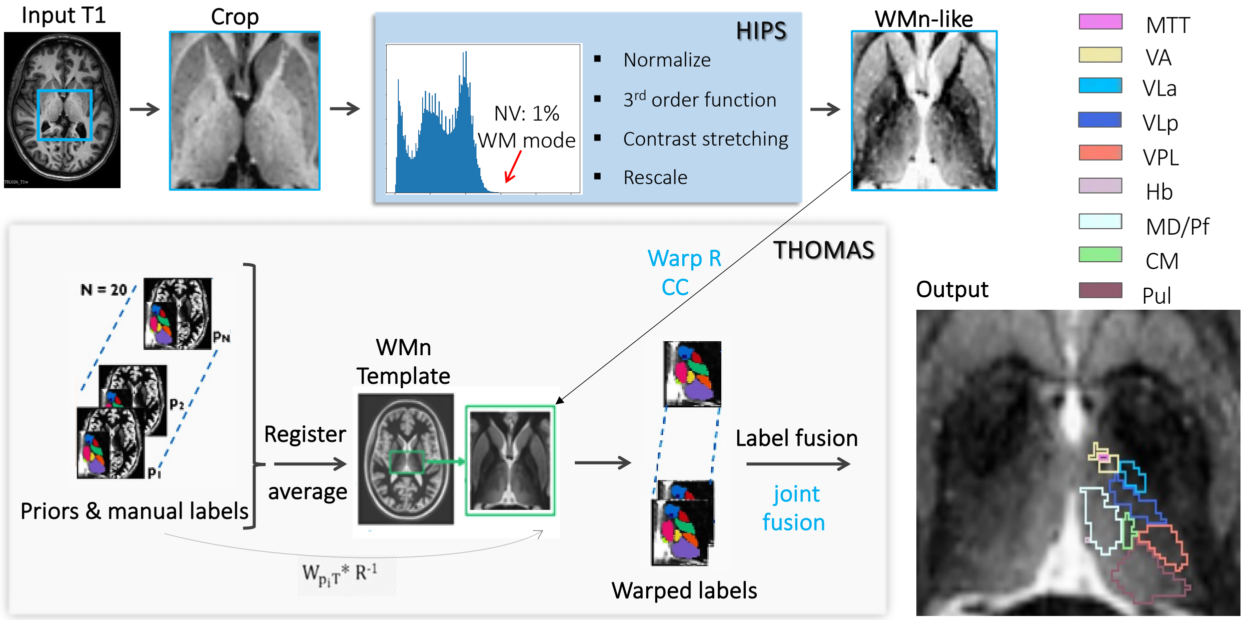


Supplemental Figure 1 – Overview of the HIPS-THOMAS methods for automated segmentation of thalamic nuclear regions.

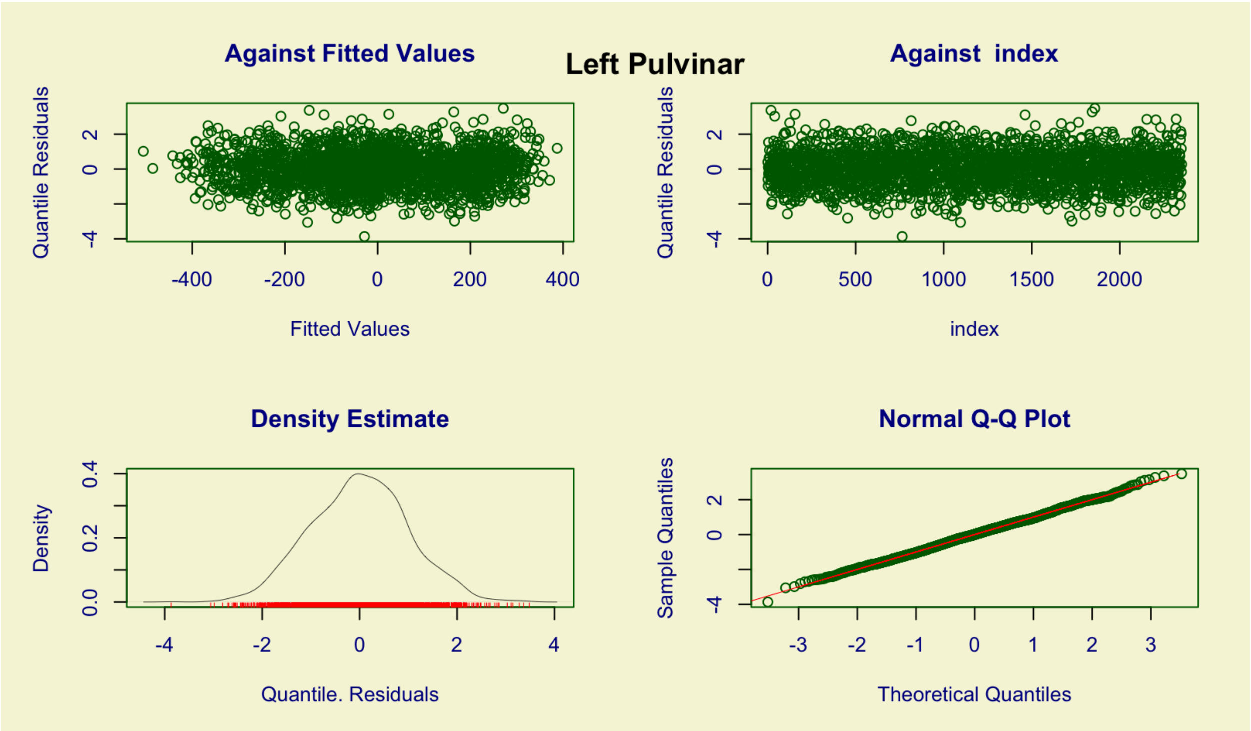


Supplemental Figure 2 – Representative model calibration plot for the left Pulvinar.

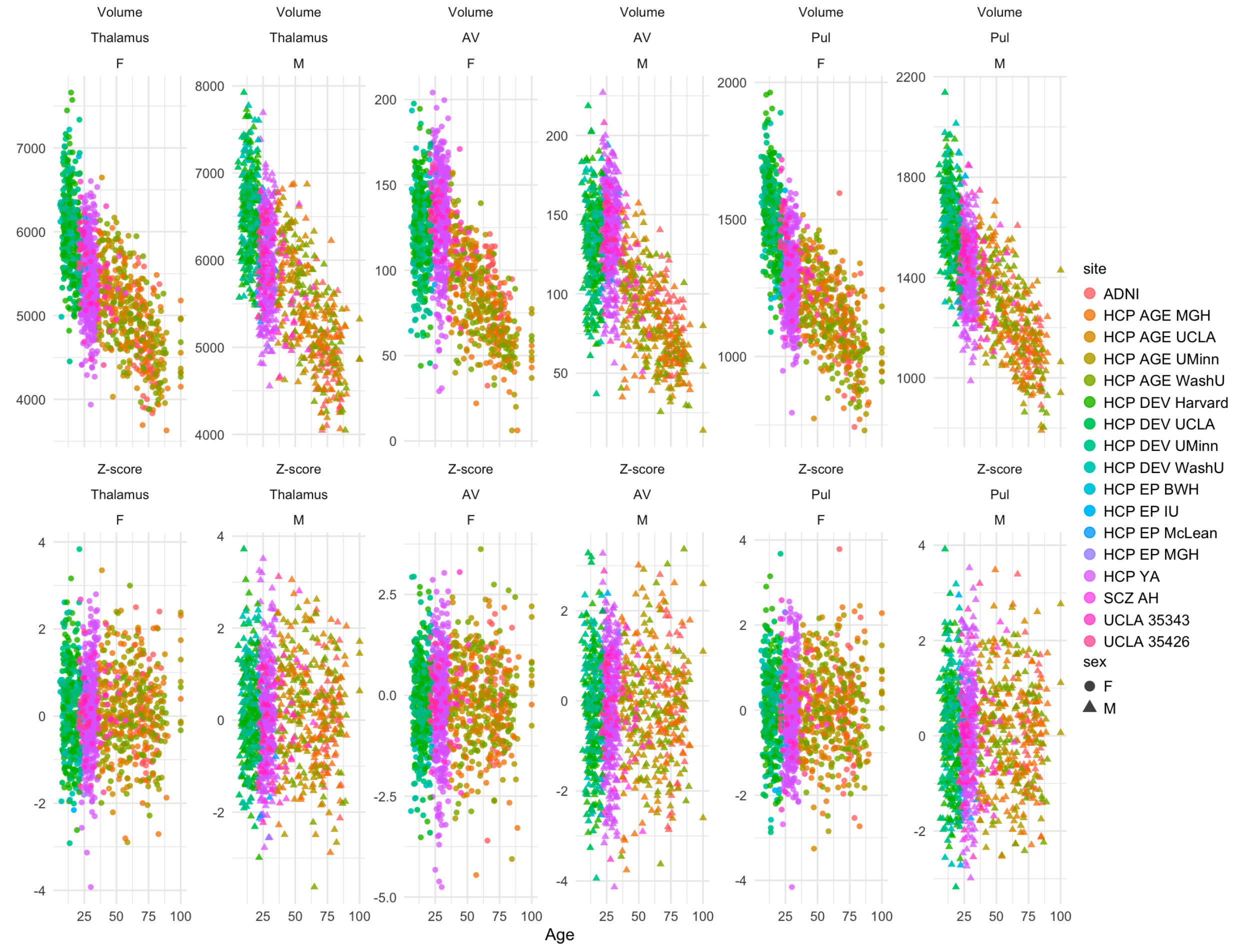


Supplemental Figure 3 – Raw volumes and z-scores for representative regions generated using the multivariate GAMLSS model.

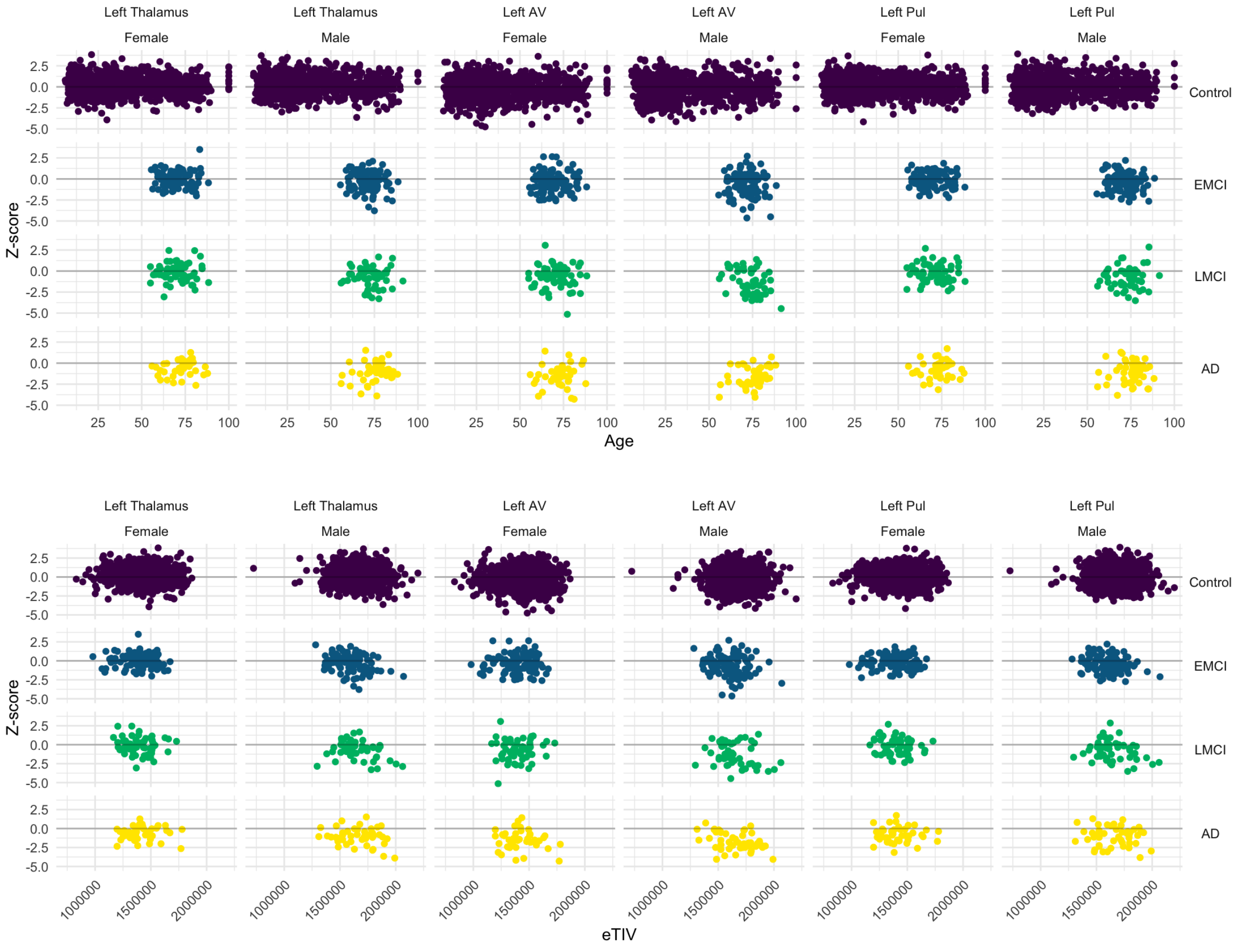


Supplemental Figure 4 – Z-scores for control subjects plotted against age (top) and eTIV (bottom) for representative regions and sex.

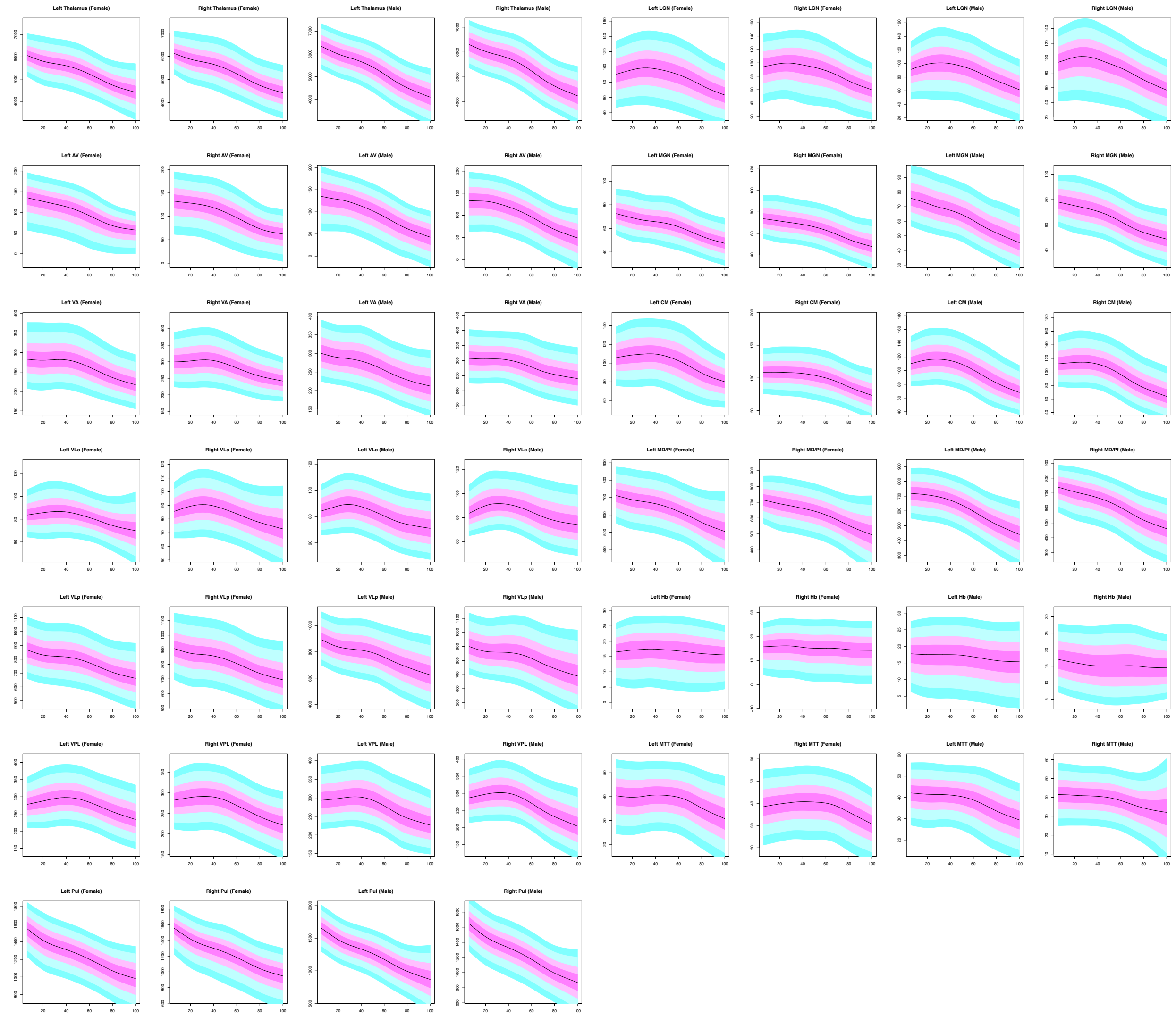


Supplemental Figure 5 - Centile plots of age versus eTIV adjusted volumes for all thalamic regions in sex stratified datasets (i.e. a univariate model). Combat has been applied to mitigate site effects.

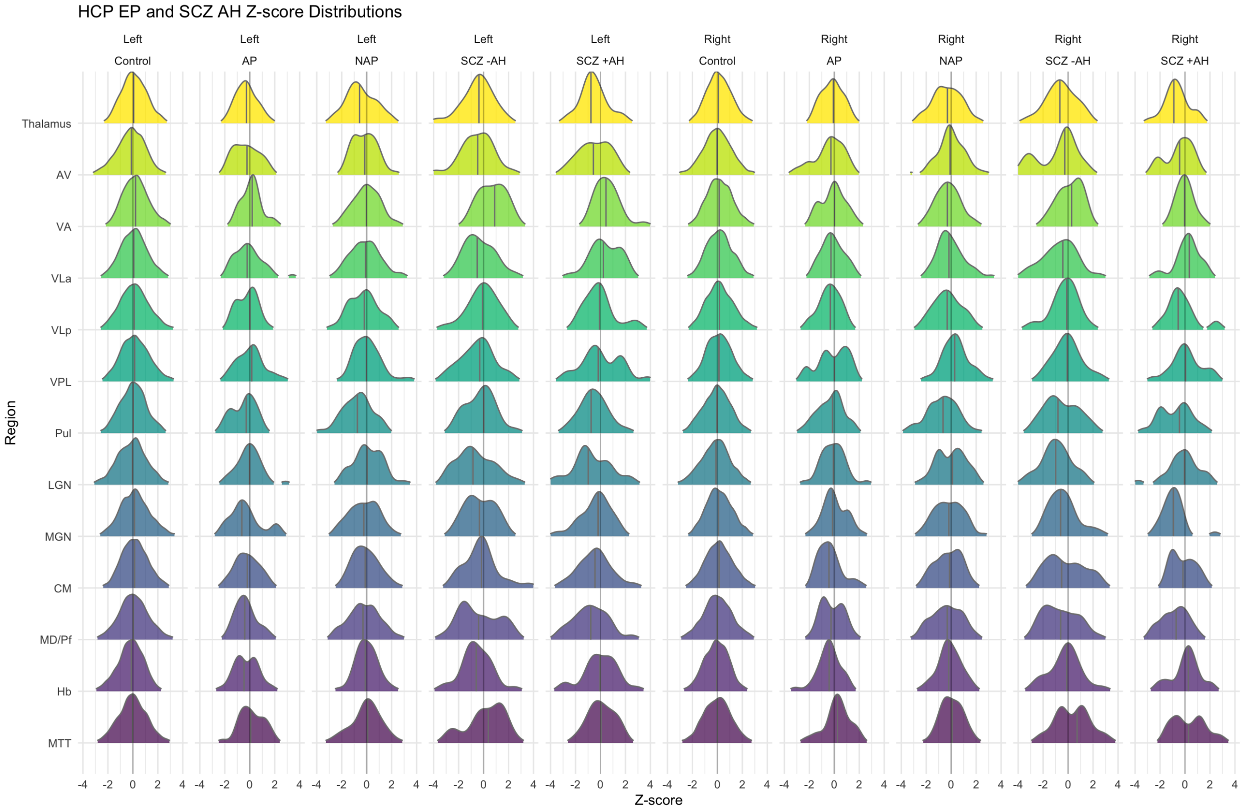


Supplemental Figure 6 - Z-score distributions for the HCP EP and SCH AH studies. Controls represent the larger pool of control subjects included in the normative model.

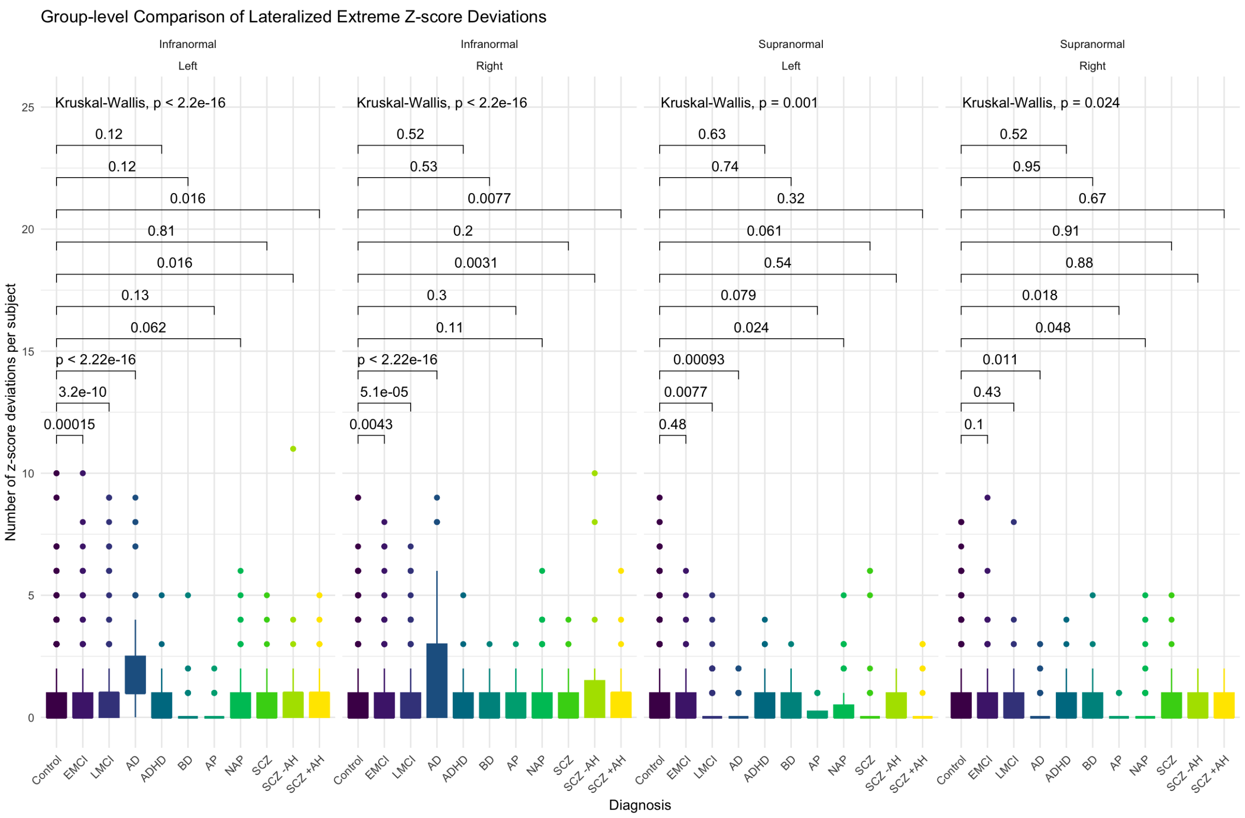


Supplemental Figure 7 – Group-level comparison of lateralized extreme z-score deviations between cases and controls.

### Supplemental Tables

Supplemental Table 1 - Average training time in seconds for each model when trained on the left whole thalamus.

| Model | Mean | Standard Deviation |
|---|---|---|
| OLS Univariate | 0.00080 | 0.00015 |
| OLS Multivariate | 0.00335 | 0.00469 |
| MFP Univariate | 0.03940 | 0.00801 |
| MFP Multivariate | 0.20631 | 0.01024 |
| GAMLSS Univariate | 5.93871 | 0.09689 |
| GAMLSS Multivariate | 14.37841 | 0.15468 |

Supplemental Table 2 – The percentage of individuals in each phenotype with at least 1 extreme z-score deviation.

| | Percentage With | | Percentage Without | |
|---|---|---|---|---|
| Phenotype | Infranormal | Supranormal | Infranormal | Supranormal |
| Control | 42.4 | 48.4 | 57.6 | 51.6 |

| | | | | |
|---|---|---|---|---|
| EMCI | 52.1 | 52.6 | 47.9 | 47.4 |
| LMCI | 67.3 | 43.4 | 32.7 | 56.6 |
| AD | 82.8 | 33.3 | 17.2 | 66.7 |
| ADHD | 52.5 | 50.0 | 47.5 | 50.0 |
| BD | 41.3 | 50.0 | 58.7 | 50.0 |
| AP | 39.3 | 25.0 | 60.7 | 75.0 |
| NAP | 49.4 | 34.9 | 50.6 | 65.1 |
| SCZ | 41.5 | 41.5 | 58.5 | 58.5 |
| SCZ -AH | 65.2 | 52.2 | 34.8 | 47.8 |
| SCZ +AH | 61.9 | 42.9 | 38.1 | 57.1 |
